# The Gut Microbiome of Youth Who Have Behavioral and Mental Health Problems: A Scoping Review

**DOI:** 10.1101/2021.09.10.21263409

**Authors:** Cherry Y. Leung, Sandra J. Weiss

## Abstract

**Background:** Mental health conditions have increased over the past several decades. While there is growing evidence that the gut microbiome affects mental health, there are limited studies focused on children, adolescents, and young adults. This scoping review examined the existing literature and compared findings on the relationships between the gut microbiome with mental health across these younger age groups.

**Methods:** A literature search using PubMed, PsycINFO, and CINAHL was performed, and bibliographies were manually searched. Eighteen articles met eligibility for our scoping review. Findings from each study were evaluated, focusing on bacterial composition and diversity among children/adolescents and young adults.

**Results:** There were no studies specifically on the adolescent age group, so data was synthesized comparing the child/adolescent (2 to <18 years of age) and young adult (18-25 years of age) groups. Studies utilized several different methods for gut microbiome analysis and examined various mental health conditions. Findings for both age groups were mostly inconsistent. However, *Bifidobacteria* seems to be associated with better mental health. Alpha diversity was lower for children/adolescents with ADHD and high stress but higher for young adults with ADHD and Major Depressive Disorder.

**Limitations:** There were inconsistencies across studies, likely due to differences in mental health problems examined, populations assessed, and research designs or measurements used.

**Conclusions:** Future research should replicate studies to confirm findings, examine lower taxonomic levels, consider longitudinal designs to assess for directionality, and consider clinical trials to examine the effects of probiotics with the same strains to manage mental health symptoms.

## Introduction

Mental illness is a major health concern throughout the world, where an estimated 792 million people (10.7%) live with a mental health disorder.^1^ Behavioral and mental health problems can manifest during childhood and early adolescence, with the prevalence increasing throughout adolescence, and the risk of recurrence during adulthood.^2,3^ Behavioral and mental health problems among youth are also predictors of many high-risk behaviors, such as risky sexual behavior,^4^ suicide,^5^ drug and alcohol abuse,^6^ and cigarette smoking.^7^ Mental health issues may also lead to increased health care utilization due to their effects on physical health, thereby having a toll on the individual as well as an economic impact.^8^

The human brain undergoes significant remodeling during childhood and adolescence. Brain development, specifically the prefrontal cortex, the region that controls and regulates executive functions, continues a wiring and rewiring process until 25 years of age.^9,10^ Thus, there is a growing acceptance that adolescence is an extended period from 10 through 25 years of age. These dynamic changes in the brain, accompanied by socioenvironmental influences and hormonal impacts of puberty, put youth at higher risk for behavioral and mental health conditions during the period from childhood to young adulthood.

There have been rapid developments and interest in determining the biological risks of mental health conditions, from predisposing genotypes to genetic alterations of our gut microbiota. The term gut microbiome describes the dynamic collection of microorganisms such as bacteria, viruses, protozoa, and fungi, and their collective genetic material that are present in the large intestine of the gastrointestinal (GI) tract.^11^ The gut microbiome is reported to have 100 times more genes than the human genome, with 10^13^ microorganisms, over 1000 unique bacterial species, and as many as 22 million unique genes.^12^ The gut microbiome has been associated with several aspects of physical health, including obesity,^13^ and immune-mediated disorders including asthma, inflammatory bowel disease (IBD), type 1 diabetes, and multiple sclerosis.^14^ Gut microbiota composition and function act as potential biomarkers of gut-brain-axis activity, playing a potential role in the pathogenesis and development of behavioral and mental health of individuals. The gut-brain-axis is a bi-directional organized system of biochemical signaling between the nervous system and enteric nervous system of animals.^15^ The gut microbiota is associated with the brain and behavior via neural, metabolic, endocrine, and immune pathways,^16^ with healthy gut function linked to normal central nervous system function.

The gut microbiota is believed to undergo significant change throughout infancy and childhood. The composition of the gut microbiota is highly variable in infancy. Coupled with anaerobic conditions created by the gut from the consumption of oxygen by facultative bacteria, ingestion of human milk oligosaccharides in breast milk results in a gut composed primarily of the genera *Bacteroides, Bifidobacterium*, and *Clostridium*.^17-19^ Some studies suggest that the core gut microbiota is established by the end of infancy, such that the GI tract of both children and adults consists predominantly of the phyla *Bacteroidetes* and *Firmicutes*.^20,21^ However, recent metagenomic studies suggest that the gut microbiota of children and adolescents differ significantly from that of adults.^22^ For example, Agans et al^22^ found that adolescents had a higher abundance of *Bifidobacterium* and *Clostridium* as compared to adults. Another study examining pre-adolescents found that children’s GI tract was enriched with *Bifidobacterium* species (spp.) and *Faecalibacterium* spp., whereas the adult gut microbiota had more *Bacteroides* spp.^23^

Furthermore, dysbiosis and inflammation of the gut have been linked to several mental illnesses, whereas probiotic administration can lead to the restoration of normal microbial balance.^24^ Probiotic research has additionally provided promising findings that manipulation of the gut microbiota has a potential role in treatment and prevention of some mental health disorders, such as depression and anxiety.^25^ However, there has been no synthesis of findings from studies examining potential links between gut microbiota composition or diversity and behavioral and mental health of children and youth. Therefore, this scoping review aims to identify existing knowledge regarding associations of the gut microbiota to behavioral and mental health of youth ages 2 to 25 years of age.

## Methods

The scoping review was undertaken by a research team including 2 mental health researchers with clinical expertise in treating mental health conditions throughout the lifespan. We utilized the framework for scoping reviews by Arksey and O’Malley^26^ to conduct this search. The review included five steps: 1) identifying the research question; 2) identifying relevant studies; 3) selecting the studies; 4) charting the data (recording significant study findings of each publication); and 5) collating, summarizing, and reporting the results.

### Research question

Our review was guided by two research questions: 1) How are behavioral and mental health problems of youth related to the composition and diversity of their gut microbiome? 2) What are the potential differences in these associations among children, adolescents, and young adults?

### Search strategy

A literature search using PubMed, PsycINFO, and CINAHL was performed to include peer-reviewed articles published prior to June 1, 2021 (see Table 1). Additional relevant articles were sought through a manual bibliography search. There was no restriction on the type of study design.

**Table 1.** Search strategy

| Construct | Search terms |
| --- | --- |
| Gut microbiome | ("microbiome" OR "flora" OR "microbes" OR "microbiota" OR "dysbiosis" OR "gut" OR "stool" OR "fecal" OR "probiotic" OR "prebiotic" OR "synbiotic" OR "psychobiotic") |
| Behavioral and mental health | AND ("mental health" OR "mental illness" OR "behavioral health" OR "neurodevelopment" OR "behavioral problem" OR "behavioral problems" OR "neurodevelopment disorders" OR "emotional problem" OR "emotional problems" OR "externalizing" OR "internalizing" OR "SMI") |
| Child | AND ("child" OR "children" OR "pediatric" OR "pediatrics" OR "preschool" OR "toddler" OR "kindergarten" OR "elementary school" OR "primary school") |
| Adolescent | OR ("adolescent" OR "adolescence" OR "teen" OR "teens" OR "teenager" OR "teenagers" OR "youth" OR "youths" OR "secondary school" OR "middle school" OR "high school") |
| Young adult | OR ("young adult" OR "college") |

Two raters agreed on the search terms. Initially, CYL screened titles to determine eligibility. Abstracts and full texts were then independently screened by CYL, and SJW reviewed the remaining full texts for eligibility and came to a consensus about the inclusion or exclusion of the article.

### Eligibility criteria

Inclusion criteria were: 1) articles in English; 2) articles where behavioral and mental health and gut microbiome were an independent variable, dependent variable, or covariate; 3) behavioral and mental health outcomes included diagnosis and symptomology; and 4) participants between the ages of 2 and 25. If adults were included, the majority of participants were between 2 and 25 years of age or the mean age was between 2 to 25 years of age.

Articles were excluded if: 1) the study was primarily focused on the adult population over 25 and the mean age was over 25 years; 2) population range was not specified; 3) was not an original study (ie., case report, review, commentary, letter to the editor, or erratum); 4) the study investigated non-human animals; and 5) the study lacked measurement of the gut microbiome.

### Screening of publications

The search yielded 1,623 articles of which 289 duplicates were removed. Of the remaining, 1,098 articles were excluded based on the title or abstract. Of the total number of studies included for full-text review, 88 were included. Of these remaining articles assessed for eligibility, 12 were included. An additional 6 articles were included from a manual search of the bibliographies of the remaining 12 articles. These procedures resulted in a total of 18 articles being eligible for this scoping review. A table was developed to extract the characteristics of each study to include the author and year of publication, study design, sample size, age of participants, sample characteristics, assessments utilized, sequencing of stool samples, and results (see Table 2).

**Table 2.** Gut microbiome and behavioral and mental health studies used in the included articles and their features

| Publication | Country | Study design | Sample size (n) | Age (years) | Sample characteristics | Behavioral and mental health focus | Microbiome analysis | Results |
| --- | --- | --- | --- | --- | --- | --- | --- | --- |
| Aarts et al<sup>44</sup> | Netherlands | Case-control | 96 | ADHD: 19.5 (SD = 2.5)<br>HC: 27.1 (SD = 14.3) | ADHD participants were from the NeuroIMAGE study and dx. was based on DSM-IV using the SADSS. Healthy controls were either healthy participants and unaffected siblings of individuals with ADHD from the NeuroIMAGE study or self-reported healthy volunteers from the Brain Imaging Genetics study. | ADHD<br>Assessments: fMRI analysis using reward anticipation | 16S rRNA (V3-V4 region), 454 Life Sciences GS-FLX platform; presence of KEGG Orthogs and subsequent functional and metabolic pathways using PICRUST | Composition: ADHD participants had more Actinobacteria as well as *Bifidobacterium*, which is linked to the encoding of cyclohexadienyl dehydratase, compared to controls. Diversity: Mean Shannon Index 5.3 for ADHD group and 5.2 for control group; Mean Chao Index 604.5 for ADHD group and 579.7 group. Other: Independent of ADHD diagnosis and age, increased functionality of cyclohexadienyl dehydratase was associated with decreased ventral striatal fMRI responses during reward anticipation. |
| Flannery et al<sup>30</sup> | USA | Cross-sectional | 40 | 5-7<br>M = 6.12 (SD = 0.69) | Participants were a subsample of families from a larger study conducted in the Stress Neurobiology and Prevention laboratory. | Behavior<br>Assessments: CBQ and CBCL for internalizing (depression and anxiety) and externalizing (inhibitory control, aggression) symptoms, LEC for socioeconomic risk, socioeconomic status, and adverse home environment exposure, PSI, IEMP, and FFMQ for perceived parental stress and well being | Shotgun metagenomics | Composition: Socioeconomic risk exposure and child behaviors were associated with the relative abundances of *Bacteroides* and *Bifidobacterium* spp. and functional modules encoded in their genomes (monoamine metabolism) Diversity: There were no associations between behavioral dysregulation and caregiver behavior and gut diversity. |
| He et al<sup>41</sup> | China | Longitudinal prospective observational | HR: 81<br>UHR: 19<br>HC: 69 | 13-30<br>HR: M = 21.67 (SD = 5.75)<br>URH: M = 20.47 (SD = 4.57)<br>HC: M = 23.13 (SD = 3.89) | HR: One first-degree relative with SCZ<br>UHR: Screened with SIPS, had one of the following: BIPS, APSS, or GRDS<br>HC: No first-degree relative with SCZ<br>Exclusion: GI and endocrine diseases, serious organ disorders, hx of psychiatric disorder dx and treatment, and alcohol, antibiotic, and probiotics in the last 3 months | Psychosis<br>DSM-IV, SOPS for prodromal sx, GAF-M for global functioning | 16S rRNA (V4 region, 515F/806R), Illumina MiSeq 250 bp paired-end | Composition: UHR, compared to HR and HC, individuals had higher levels of *Clostridiales*, *Lactobacillales*, and *Bacteriodales*. Differences in gut microbiota composition are related to increased SCFA production. Diversity: There were no differences with alpha diversity, as measured with Shannon Index, among the HR, UHR, and HC. For beta diversity as measured with Canberra distance matrix, there were group differences between UHRs and HRs with HCs. |
| Ishii et al.<sup>33</sup> | Japan | Cross-sectional | OI: 56<br>HC: 9 | 8.8-17.2<br>OI: M = 13.6<br>(SD = 1.6)<br>HC: M = 12.3<br>(SD = 3.8) | Study focused on patients with OI, who displayed depressive and anxiety symptoms. Exclusion: Patients with other underlying diseases, such as IBD, infectious enteritis, and eating disorders, and no antibiotics or probiotics for 2 weeks prior to testing. | Internalizing and emotional<br>Assessments: OI diagnosed with Japanese Clinical Guidelines for Juvenile Orthostatic Dysregulation Version 1<sup>2</sup>, Japanese version of CDI for depression and CMAS for anxiety | 16S rRNA | Composition: OI patients had higher proportion of Clostridium subcluster XiVa and/or Enterobacteriaceae than HC. Among OI, Bifidobacterium was less frequent in the depressed group, compared to non-depressed. Diversity: There were no differences in the gut diversity of 1) controls, depressed, and nondepressed groups and 2) anxiety and nonanxiety groups. |
| Jiang et al.<sup>40</sup> | China | Cross-sectional | A-MDD: 29<br>R-MDD: 17<br>HC: 30 | 18-40<br>A-MDD: M = 25.3 (SD = 5.4)<br>R-MDD: M = 27.1 (SD = 5.4)<br>HC: M = 26.8 (SD = 5.4) | A-MDD participants had HAMDS score ≥20. R-MDD has a baseline HAMDS score ≥20 upon admission to the hospital. Exclusion: hypertension, cardiovascular disease, diabetes mellitus, obesity, liver cirrhosis, fatty liver disease, IBS, IBD, drug or alcohol abuse in last year, antibiotic, probiotic, prebiotic, and symbiotic use one more before fecal sample collection, active bacterial, fungal, or viral infections. | Depression<br>A psychiatrist screened participants with the Mini-International Neuropsychiatric Interview. SCID for screening major depressive disorder, HAMDS and MADRS for depressive symptom severity | 16S rRNA (V1-V3 region), Roche 454 FLX system 600 bp | Composition: A-MDD and R-MDD groups had increased Bacteroidetes, Proteobacteria, and Actinobacteria, as well as reduced Firmicutes compared to HC. MDD groups had increased levels of Enterobacteriaceae and Alistipes and reduced levels of Faecalibacterium. Faecalibacterium was negatively correlated with depressive symptom severity. Diversity: Increased alpha diversity, as measured with Shannon Index, was reported in the A-MDD group, compared to HCs. Clusters among the three groups could not be created to estimate beta diversity because of significant interindividual variation. |
| Jiang et al.<sup>36</sup> | China | Cross-sectional | ADHD: 51<br>HC: 32 | 6-10<br>ADHD: M = 8.47<br>HC: M = 8.5 | Severity of ADHD symptoms were assessed in treatment naïve children. Exclusion: probiotics and antibiotics 2 months prior to stool sample collection, apparent GI symptoms, depressive and anxiety symptoms, obesity, common childhood atopic diseases, and hx of current use of ADHD medications. | ADHD<br>Assessments: CPRS for parental assessment of ADHD symptom severity | 16S rRNA (V3-V4 region, 338F/806R), Illumina MiSeq | Composition: Compared to HC, the ADHD group had a decrease in Faecalibacterium. Among ADHD, Faecalibacterium abundance was negatively associated with parental reports of ADHD symptoms. Diversity: There were no differences in alpha diversity between ADHD and HC groups. Due to significant interindividual variation, beta diversity could not be discriminated between the two groups. |
| Kato-Kataoka et al.<sup>38</sup> | Japan | Double-blind, placebo-controlled, parallel-group | Tx: 24<br>Placebo: 23 | Tx: M = 22.8 (SD = 0.4)<br>Placebo: | Participants were healthy medical students. Tx group took strain Shirota fermented milk and placebo | Anxiety and stress<br>STAI for state and trait anxiety levels, NEO- | 16S rRNA (V1-V2 region), Illumina MiSeq. LLV agar | Composition: 16S rRNA showed a lower percentage of Bacteroidaceae in tx group. Diversity: Phylogenetic diversity |
|  |  | clinical trial |  | M = 22.8)<br>SD = 0.3) | milk daily for 8 weeks. Exclusion: Habitual smoking, ≥30 years, high scores on STAI-trait and GHQ-28. | FFI for five personality traits (neuroticism, extraversion, openness, agreeableness, and conscientiousness) at time of screening, VAS for survey of stress, saliva for cortisol and alpha-amylase, peripheral venous blood for NK cell activity | medium and colony PCR to identify living fecal *L. casei* strain Shirota CFU | was higher among the tx group, compared to the placebo group before examination. The Shannon Index of the placebo group was decreased pre-examination compared to staying constant in the tx group. Additional findings: Tx group had less feelings of stress, abdominal function, and fewer changes in leukocyte gene expression compared to the placebo group. Salivary cortisol levels before the examination (stressor) was increased in the placebo group. Tx group had reduced GI symptoms. |
| Loughman et al<sup>29</sup> | Australia | Longitudinal cohort | 201 | 2 when behavioral outcome measured | Sub-cohort from the Barwon Infant Study with infants who had stool samples at 1, 6, and 12 months of age. | Behavior<br>Assessments: CBCL for child behavior, including internalizing, externalizing, and total problems subscales, at 2 years old, infant temperament at 1, 6, and 12 months measured by parent report | 16S rRNA (V4 region), Illumina MiSeq | Composition: Decreased abundance of *Prevotella* at 12 months was associated with increased behavioral problems at 2 years, especially in internalizing problem scores. Antibiotic use was the best predictor of decreased *Prevotella*. Diversity: There were no associations between alpha diversity or beta diversity with behavioral outcomes at 2. |
| Loughman et al<sup>34</sup> | Australia | RCT and longitudinal | 118 | 2 when behavioral outcome measured, Initial enrollment M = 7.4 weeks (SD = 2.7 weeks) | Infant with colic less than 3 months older were enrolled in a 4-week RCT of Lactobacillus reuteri DSM 17938 probiotic supplement and 2 year follow-up behavioral data were included. | Behavior<br>Assessments: CBCL for child behavior, including internalizing, externalizing, and total problems subscales, at 2 years old, Wessel's criteria for parent report of presence of colic at study enrolment | 16S rRNA (V3-V4 region), Illumina MiSeq 2x300 bp paired-end | Composition: *Bifidobacterium*, *Clostridium*, *Lactobacillus*, and *Klebsiella* were associated with colic severity, while baseline microbiota composition predicted further crying at 4 week with up to 65% accuracy. Diversity: Alpha diversity of fecal microbiota at baseline collection was associated with birth mode, feed type, and child gender, but not crying and behavioral outcomes. |
| Ma et al<sup>42</sup> | China | Cross-sectional, exploratory | FSCZ: 40<br>TSCZ: 85<br>HC: 69 | SCZ: M = 24.19 (SD = 6.18)<br>HC: M = 23.14 (SD = 3.2) | FSCZ group had less than ≤12 months SCZ dx and no treatment with antipsychotic medications or other psychotropics. TSCZ group had ≥12 months dx and received antipsychotic treatment ≥3 months. HC group had no physical and mental health disorders. Inclusion: 16-40 years old, normal BMI, right-handed, not antibiotics, probiotics, or prebiotics within one month prior to stool collection. | Psychosis<br>Structured Clinical Interview for DSM IV Disorders Criteria for diagnosis, PANSS for psychotic symptom severity, Structural MRI for T1-weighted brain imaging data. | 16S rRNA (V4 region, 515F/806R), 250bp reads | Composition: FSCZ and TSCZ had changes for the families *Christensenellaceae*, *Enterobacteriaceae*, *Pasteurellaceae*, *Turicibacteraceae* and genus *Escherichia* compared to HC. Specific SCZ-associated microbiota were correlated with changes in right middle frontal gyrus volume of SCZ patients. Diversity: TSCZ compared to HC had low microbiome alpha diversity indexes. |
| Michels et al<sup>32</sup> | Netherlands | Cross-sectional | 93 | 8-16 | Participants were Dutch-speaking Belgian children from the ChiBS study. An extra study wave between March-May 2015 was done to collect stool samples of 8-16 year olds. Exclusion: Heart and autoimmune diseases, oral anti-inflammatory drugs and SSRIs, oral corticosteroids and antibiotics 3 months prior to stool collection. | Internalizing and emotional<br>Assessments: Hair cortisol for chronic stress exposure, HRV for parasympathetic activity, the CLES-C for significant life events, self-report of negative and positive emotions, and parent-reported SDQ for emotional problems. | 16S rRNA (V3-V4 region), Illumina MiSeq 300 bp paired-end | Composition: High stress associated with lower Firmicutes at the phylum level and higher *Bacteroides*, *Parabacteroides*, *Rhodococcus*, *Methanobrevibacter*, and *Roseburia* but lower *Phascolarctobacterium* at the genus level. Conflicting results reported for different stress measures. Different results reported between pre-adolescents and adolescents. Diversity: High stress was associated with lower alpha diversity, and specifically among preadolescents but not adolescents. Beta diversity was distinct among the high and low stress groups within HRV and happiness measurements, and only among preadolescents. |
| Michels et al<sup>27</sup> | Netherlands | Cross-sectional | 113 | 8-16<br>M = 12.7<br>(SD = 1.7) | Participants were Dutch-speaking Belgian children from the ChiBS study. An extra study wave between March-May 2015 was done to collect stool samples of 8-16 year olds. Exclusion: Heart and autoimmune diseases, oral anti-inflammatory drugs and SSRIs, oral corticosteroids and antibiotics 3 months prior to stool collection. | Internalizing and emotional<br>Assessments: Hair cortisol for chronic stress exposure, HRV for parasympathetic activity, the CLES-C for significant life events, self-report of negative and positive emotions, and parent-reported SDQ for emotional problems. | Short-chain fatty acid analysis, gas chromatography, analysis of fecal calprotectin, a marker of gut mucosal inflammation | Composition: N/a. Diversity: N/a. Additional Findings: Emotional problems were associated with higher butyrate, valerate, isovalerate, and isobutyrate. HRV was related to lower valerate levels. Hair cortisol and fecal calprotectin were not associated with short-chain fatty acids. Stress measures were not associated with fecal calprotectin. |
| Nishida et al<sup>39</sup> | Japan | Double-blind, placebo-controlled, parallel-group clinical trial | CP2305: 29<br>Placebo: 31 | CP2305: M = 24.9<br>(SD = 0.5)<br>M = 25.3<br>(SD = 0.6) | Participants were sixth year medical students who were randomly allocated to the *Lactobacillus gasseri* CP2305 or placebo group. Participants took 2 tabs (placebo or CP2305) once daily for 24 weeks. Exclusion: medications that affect sleep quality, hormonal contraceptives, and active disease (mental, inflammatory, bone, and hormonal disorders), medications 3 months prior to study enrollment. | Anxiety and stress<br>STAI, GHQ-28, and HADS for mental health, PSQI for sleep, salivary cortisol for stress levels, and EEG for overnight brain activity. | 16S rRNA (V2 region), Illumina MiSeq. SCFA concentration measurement | Composition: CP2305 intake reduced anxiety and sleep disturbance compared to placebo. CP2305 administration attenuated the stress-induced decline of *Bifidobacterium* spp. and stress-induced elevation of *Streptococcus* spp. Diversity: N/a. |
| Prehn-Kristensen et al<sup>28</sup> | Germany | Cross-sectional | ADHD: 14<br>HC: 17 | 6-16<br>ADHD: M = 11.9<br>(SD = 2.5)<br>HC: M = 13.1<br>(SD = 1.7) | Children and their parents were interviewed in German by experienced child and adolescent psychiatrists and psychologists. 6 ADHD patients also met criteria for ODD. 10 ADHD patients were taking ADHD medications for more than 1 year, 9 of which stopped taking the medicine for at least 48 hours prior to stool sample collection. | ADHD<br>Assessments: K-SADS-PL for psychopathology, CBCL for behavior, internalizing and externalizing problems, FBB-HKS for ADHD rating scale, WURS-K for ADHD symptoms, and ADHD-SB for ADHD rating scale of adults (parents) | 16S rDNA (V1-V2 region), Illumina MiSeq 200bp | Composition: ADHD patients had higher levels of *Bacteroidaceae*, *Neisseriaceae* and *Neisseria species*. Diversity: Alpha diversity was decreased and beta diversity differed in ADHD patients compared to HC. |
| Schwarz et al<sup>44</sup> | Finland | Case control and longitudinal | FEP: 28<br>HC: 16 | 18-40<br>FEP: 25.9<br>(SD = 5.5)<br>HC: 27.1<br>(SD = 6.0) | Inclusion: Primary psychotic disorders<br>Exclusion: Substance-induced psychoses and psychotic disorders due to a general medical condition<br>HC: matched controls | Psychosis<br>DSM-IV, BPRS-E and SANS for positive and negative sx, GAF for global functioning, medical hx, Health Behavior and Health among the Finnish Adult Population survey for diet, and Gothenburg scale for physical activity | qPCR for 16S primers, Illumina Hi Seq 2000 100 bp paired-end, analysis of 7 bacterial groups: lachnospiraceae, Ruminococcaeae, *Bacteroides* spp., *Atropobium*, *Bifidobacteria*, and *Lactobacillus* groups | Composition: *Lactobacillaceae* differed between FEP individuals and HC. *Lactobacillus* had a positive correlation with psychotic sx severity and negative correlation with global assessment of functioning. FEP individuals with strongest microbiota differences had poorer response to antipsychotic treatment at 12 months. Diversity: N/a. |
| Stevens et al<sup>35</sup> | New Zealand | Double-blind, RCT | 17 | 7-12<br>Treatment: M = 9.39<br>(SD = 2.9)<br>Control: M = 9.3<br>(SD = 1.3) | Male participants were from a sub-cohort of a larger study examining dietary micronutrient supplementation on ADHD outcomes. The micronutrient (capsule with blend of vitamins, minerals, amino acids, and antioxidants) group and placebo group had pre-RCT and post-RCT stool samples and were treated for 10 weeks. Exclusion: probiotics at time of study | ADHD<br>Assessments: CGAS for overall level of functioning, ADHD-RS-IV for ADHD symptoms, and a brief diet intake questionnaire at baseline and end of the RCT. | 16S rRNA (V3-V4 region), Illumina MiSeq 220bp | Composition: The micronutrient tx group did not have large-scale changes in composition and structure of the microbiome. *Bifidobacterium* was decreased and *Collinsella* was increased in the post-micronutrient tx group. Decreased Actinobacteria was correlated with decreased ADHD-IV-RS scores. Diversity: Observed OTUs in the placebo and tx groups were different, and the placebo group had a significant decrease in alpha diversity between pre- and post-RCT. There were no significant differences among beta diversity pre- and post-RCT for either group. |
| Wang et al<sup>37</sup> | Taiwan | Cross-sectional | ADHD: 30<br>HC: 30 | 6-16<br>ADHD: M = 8.4<br>HC: M = 9.3 | ADHD medication naïve participants who were clinically diagnosed by a psychiatrist. Exclusion: hx of neuropsychiatric diseases or | ADHD<br>Assessments: K-SADS-E to assess current and past episodes of | 16S rRNA (V3-V4 region), Illumina MiSeq 550bp | Composition: ADHD group had decreased *Bacteroides coprocol* and increased *Bacteroides uniformis*, *Bacteroides ovatus*, and *Sutterella stercoricanis*. *B.* |
|  |  |  |  |  | major physical illnesses and vegetarians or were currently taking probiotics or antibiotics | psychopathology of children and teenagers, ADHD-RS for ADHD symptoms, and FFQ to evaluate dietary intake within the previous year. |  | *ovatus* and *S. stercoricanis* were positively correlated with more ADHD symptoms. *S. stercoricanis* was associated with dairy, nuts/seeds/legumes, ferritin, and magnesium. Diversity: ADHD patients had higher Shannon Index and Chao Index than HC, while the Simpson index was lower for the ADHD group compared to HC. Gut microbiota communities were similar among the two groups. |
| Yuan et al<sup>43</sup> | China | Longitudinal, intervention | FESCZ:41<br>HC: 41 | FESCZ: 23.1<br>(SD = 8.0)<br>HC: 24.7<br>(SD = 6.7) | FESCZ: Drug-naïve first-episode SCZ, normal weight patients enrolled and given 24-week risperidone tx<br>Inclusion: PANSS score >60, normal vaginal delivery birth, normal body weight<br>HC: No previous hx of psychiatric diseases, not matched<br>Exclusion: Dx with autoimmune, heart, neurological or other psychiatric diseases, pregnant or lactating women, hx of antibiotic or anti-inflammatory agent, or probiotics in past month, significant change in living environment and diet in past month, and significant diarrhea and constipation in past month | Psychosis<br>DSM-IV, PANSS for positive and negative sx, medical and psychiatric hx, physical exam, metabolic parameters (i.e., glucose, triglycerides) | qPCR for 16S primers, analysis of 5 bacterial taxa: *Bifidobacterium* spp., *Escherichia coli*, *Clostridium coccoides* group, *Lactobacillus* spp., and *Bacteroides* spp. | Composition: Compared to HC, FESCZ patients had lower numbers of *Bifidobacterium* spp., *Escherichia coli*, *Lactobacillus* spp. And higher numbers of *Clostridium coccoides* group. After 24 weeks risperidone tx, *Bifidobacterium* spp. and *E. coli* increased and *Clostridium coccoides* group and *Lactobacillus* spp. decreased.<br>Diversity: N/a. |

### Data summary and synthesis

After reviewing the table, each study was labelled according to the main behavioral and mental health focus, based on the objectives, variables used, and results of the study. For the purposes of this review, publications were classified into two age groups: child/adolescent and young adult. Children (2-12 years) and adolescents (12-18 years) were grouped into one category due to the limited studies focused solely on the adolescent age group as well as the overlap of ages in the studies.

## Results

A total of 18 published peer-reviewed articles met the inclusion criteria. Of the studies, 6 focused on children (2 to 12 years old), 4 on children and adolescents (6 to 17 years old), 1 on adolescents and young adults (13-30 years old), and 7 on young adults (>18 years old). Furthermore, of these included studies, 5 were conducted in China, 3 in Japan, 2 in Australia, 3 in the Netherlands, 1 in the USA, 1 in Finland, 1 in Germany, 1 in New Zealand, and 1 in Taiwan. All studies were quantitative, and number of participants ranged from 17 in the smallest study to 201 in the largest study. Although our search strategy included all articles published until June 2021, the articles included in this review were published within the last six years (from 2015).

The child/adolescent and young adult groups were each categorized into relevant areas based on the main behavioral or mental health focus of the research. The child/adolescent group consisted of: 1) internalizing problems, 2) externalizing problems, and 3) Attention Deficit/Hyperactivity Disorder (ADHD). Two studies reported findings on both internalizing and externalizing problems during childhood, and one study examined these problems as well as ADHD. The young adult group consisted of: 1) internalizing problems, 2) psychosis, and 3) ADHD. While the majority of studies utilized 16s rRNA sequencing to measure the gut microbiota (n=14, 78.8%), one used next generation pyrosequencing, one used 16s rDNA sequencing, one used shotgun metagenomic sequencing, while another used metabolomic analysis.

### Children/Adolescents

#### Internalizing symptoms

Six of the nine studies that focused on children/adolescents examined the associations of the gut microbiome with internalizing symptoms. Five studies were cross-sectional while the sixth was a longitudinal cohort study. The *Child Behavioral Checklist* (CBCL) was the primary assessment for measuring internalizing symptoms for four of the six studies. The CBCL is the component of the Achenbach System of Empirically Based Assessment which measures behavioral and emotional problems in children and adolescents. While three studies utilized 16s rRNA sequencing for microbiota analysis, Michels et al^27^ performed short-chain fatty acid analysis to examine metabolite composition and Prehn-Kristensen et al^28^ utilized 16S rDNA to describe bacterial composition and diversity of the gut.

Associations between diversity and bacterial dysbiosis with internalizing symptoms measured by the CBCL varied. While Loughman et al^29^ reported no differences in alpha and beta diversity of children with and without behavioral problems in their longitudinal study, they found that children with decreased *Prevotella* at 12 months had increased internalizing symptoms at two years. In a sub-cohort of infants with colic, Loughman et al^29^ reported no associations between the gut microbiota during infancy with internalizing symptoms at two years of age. Findings of five to seven year olds suggested that *Bacteroides fragilis* was associated with reduced levels of emotional reactivity, sadness, and increased inhibitory control,^30^ which correlates to lower internalizing symptoms.^31^ In a study focused on a sample between six and 16 years of age with ADHD, Prehn-Kristensen et al^28^ also examined behavioral problems using the CBCL and found no associations between gut microbiota composition and internalizing symptoms.

The two cross-sectional studies by Michels et al^27,32^ utilized the same Children’s Body Composition and Stress (ChiBS) cohort consisting of healthy Dutch-speaking Belgian children to examine the relationships between the gut microbiota and short-chain fatty acids of children between eight and 16 years of age. Both studies utilized the parent-reported Coddington Life Events Scale for Children to measure negative events that may affect the child’s health and the parent-reported Strengths and Difficulties Questionnaire subscale that measured emotional problems. Michels et al^32^ additionally measured self-reported negative emotions (sum of anger, anxiety, and sadness) and feelings of happiness. They found that happiness contributed to differences in microbiota communities between high and low stress participants in pre-adolescents but not adolescents. Those with high stress had lower Firmicutes at the phylum level, especially the order Clostridiales, and lower *Phascolarctobacterium* but higher *Bacteroides, Parabacteroides, Rhodacoccus, Methanobrevibacter*, and *Roseburia* at the genus level. Michels et al^27^ focused on short-chain fatty acids (SCFA) analysis of stool samples as each SCFA is produced by a certain set of gut microbiota and may influence appetite regulation, gut barrier protection, and inflammation. In particular, they examined fecal calprotectin, a marker of intestinal inflammation and proxy of gut barrier integrity, and SCFAs butyrate, propionate, acetate, valerate, isobutyrate, and isovalerate. Michels. et al^27^ reported that emotional problems were associated with higher butyrate, valerate, isovalerate, and isobutyrate, but there were no associations between hair cortisol and these SCFAs as well as no relationship reported between these SCFAs, stress measures, and fecal calprotectin. While Michels et al.^32^ did not explore gut microbiota diversity, they reported that high stress was associated with lower microbial diversity (alpha diversity); microbial composition (beta diversity) was different among the high and low stress groups when using heart rate variability (HRV) and happiness measurements to assess stress, and among their preadolescent subsample only.

In addition to bacterial composition differences seen in Flannery et al’s^30^ study, the butyrate-producing species *Coprococcus comes* and *Eubacterium rectale* were associated with elevated anxiety and depression as well as reduced inhibitory control while *Roseburia inulinivorans* was associated with lower depression. Due to use of shotgun metagenomic sequencing, this study was also able to examine microbial functions, which are typically grouped into two categories: functions that induce intestinal inflammation and those involved in the production of monamine precursors. Flannery et al^30^ reported that certain pathways and secretion systems (Type II, III and, IV) were significantly associated with the increase in scores for aggressive anxiety problems, anxious depression, and depressive problems.

Ishii et al^33^ measured depressive and anxiety symptoms and the gut microbiota of children with orthostatic intolerance. Their primary aim compared the gut microbiota of the orthostatic intolerance group with healthy controls and secondarily compared gut microbiota composition of depressed and non-depressed children in the orthostatic intolerance group. The depressed OI participants had less *Bifidobacterium* and no differences in bacterial diversity, compared to the non-depressed group. There were no correlations between gut microbiota composition and anxiety.

#### Externalizing symptoms

Four studies on children/adolescents reported findings on externalizing problems using the CBCL. Three of these studies reported no associations between gut microbiota composition and externalizing symptoms.^28,29,34^ However, similar to their own findings on internalizing symptoms, Flannery et al^30^ reported that *B. fragilis* was associated with reduced levels of aggressive behavior, externalizing behavior, and impulsivity. The same pathways and systems (Type II, III and, IV) identified for internalizing behavior were associated with increased scores for aggressive behavior, externalizing behavior, and anger frustration.

#### ADHD

Four of the studies focused on children/adolescents with ADHD. Three were cross-sectional studies and one was a double-blind randomized controlled trial (RCT). Assessments for ADHD symptoms varied across the studies. Three of the studies used the 16S rRNA V3-V4 region to identify bacterial composition differences between ADHD and controls and the fourth used 16S rDNA V1-V2.

Stevens et al^35^ was the only RCT within the child/adolescent gut microbiome studies. This research compared the effects of broad spectrum micronutrition administration on the gut microbiota of children with ADHD versus those in a placebo group. There were no significant changes in alpha or beta diversity pre- and post-treatment in either the micronutrient or placebo groups. However, investigators reported that the treatment group had significantly increased Operational Taxonomic Units (OTU) or groups of related bacteria; there were no changes in the placebo group. The increased trend in the level of phylum Actinobacteria (although not significant), was mostly driven by higher genus *Bifidobacterium* (p = 0.04) and was correlated with lower ADHD-IV-RS scores pre-micronutrient treatment. However, low *Bifidobacterium* abundance was associated with a low ADHD-IV-RS score post-micronutrient treatment. Lastly, the study reported no changes at the family level for Coriobacteriaceae; but at the species level, *Collinsella aerofaciens* was significantly increased in the micronutrient group (p = 0.01) although there was no correlation with ADHD-IV-RS scores.

Jiang et al^36^ reported four dominant phyla, Firmicutes, Bacteroidetes, Proteobacteria, and Actinobacteria, among both their treatment naïve ADHD and healthy control groups; but there were no major differences in alpha diversity (species richness) between these two groups. The ADHD group, compared to healthy controls, had more *Peptostreptococcaceae* and a lower percentage of *Alcaligenaceae* at the family level. At the genus level, the ADHD group, compared to healthy controls, had decreased levels of *Lachnoclostridium* and *Dialister*, and a profound decrease in *Faecalibacterium*.

Prehn-Kristensen et al^28^ reported a number of significant findings. ADHD patients had both significantly decreased alpha diversity and different beta diversity compared to controls. ADHD patients had higher levels of the family *Bacteroidaceae, Neisseriaceae*, and *Neisseria* while the controls had higher levels of *Prevotellacae*.

Wang et al’s^37^ study also found differences between alpha and beta diversity of medication naïve ADHD participants compared to healthy controls. Five phyla, including Bacteroidetes, Firmicutes, Proteobacteria, Fusobacteria, and Actinobacteria accounted for 99% of the bacteria in both groups. However, the abundance of Fusobacteria was higher in the ADHD group (median = 0.28%) compared to the healthy control group (0.02%). The bacterial profiles at the genus level were similar in the ADHD and healthy control groups, where the most abundant were *Bacteroides, Prevotella, Parabacteroides, Phascolarctobacterium*, and *Escherichia Shigella*. At the species level, the ADHD group had a decrease in the relative abundance of *Bacteroides coprocola* and increases in *Bacteroides uniformis, Bacteroides ovatus*, and *Sutterella stercoricanis*, compared to the healthy control group.

### Young Adults

#### Internalizing symptoms

Three studies reported findings on internalizing symptoms in the young adult age group. Two studies originating in Japan studied anxiety and stress levels of healthy medical students. The third study from China was cross-sectional and examined gut microbiota in a group of depressed adults. All studies used 16s rRNA sequencing to analyze the stool samples.

The two studies from Japan were clinical trials investigating the effects of probiotic administration on gut microbiota changes, cortisol levels, stress, and anxiety. Kato-Kataoka et al^38^ study administered a fermented drink containing *Lactobacillus casei* strain Shirota for 8 weeks while Nishida et al^39^ study administered *Lactobacillus gasseri* CP2305 probiotic tabs for 24 weeks. Measurements were obtained at baseline and after four weeks of probiotic administration in Kato-Kataoka et al’s^38^ study. Although healthy, these medical students were hypothesized to be under extreme stress since this study knowingly took place during their national examination to advance to the next year. In Kato-Kataoka et al’s^38^ study, the three phyla Firmicutes, Bacteroidetes, and Actinobacteria were the most abundant in both treatment and placebo groups pre- and post-treatment. Although there were no differences at baseline, the probiotic group had higher alpha diversity compared to the placebo group after experiencing stress associated with their academic examination. Additionally, the percentage of Bacteroidetes at the phylum level increased before examination in the placebo group only. Anxiety levels as measured by the State-Trait Anxiety Inventory (STAI) were normal at baseline, elevated one day before examination, and returned to baseline at two weeks after the examination; there were no significant differences seen between probiotic and placebo groups. Similarly, salivary cortisol levels increased the day before examination but only for the placebo group. There was no direct analysis of the relationship between gut microbiota changes with anxiety and stress in this study.

Nishida et al^39^ measured symptoms of anxiety and cortisol levels of fifth to sixth year medical students. They reported a reduction in anxiety symptoms as well as depression symptoms for the probiotic group but not the placebo group. They reported no differences in salivary cortisol levels between the two groups, but lower Chromoganin A levels of the probiotic group, compared to the placebo group. The placebo group had decreased *Bifidobacterium* and increased *Streptococcus* as a result of stress encountered during the national examination while the reduction of *Bifidobacterium* was mitigated and the elevation of *Streptococcus* was prevented in the probiotic group.

While the cross-sectional study by Jiang et al^40^ consisted of Chinese participants in a broader age range (18-40), the median age was between 25.3 to 27.1 years. Three groups were compared: participants with an active major depressive disorder (A-MDD) episode, those who had taken antidepressants (R-MDD; responded-MDD), and healthy controls. Most notably, both A-MDD and R-MDD, compared to healthy controls had increased levels of Enterobacteriaceae and *Alistipes* and reduced levels of *Faecalibacterium*. However, A-MDD had increased alpha diversity compared to healthy controls. There were no differences in alpha diversity between those with depression who had taken antidepressants and other groups.

#### Psychosis

Three studies were interested in the gut microbiota of patients with schizophrenia using 16S rRNA for gut microbiota sequencing, and one study examined the gut microbiota of patients with primary psychotic disorders using 16S primers. The three studies on schizophrenia were conducted in China. He et al^41^ investigated the gut microbiota differences between healthy individuals considered high-risk for developing schizophrenia based on their family history with those who had low risk. There were no significant differences in alpha diversity among the three groups. However, beta diversity was significantly different for individuals in the ultra high risk (UHR) and high risk (HR) groups compared to healthy controls. There were also increases in the orders *Clostridiales, Lactobacillales* and *Bacteroidales*, genera *Lactobacillus* and *Prevotella*, and species *Lactobacillus ruminis* for individuals in the UHR group compared to those in the HR group and healthy controls. This study additionally examined SCFA production and found that the UHR group had elevated acetyl-CoA synthesis compared to the HR group and healthy controls following adjustment for multiple tests.

Ma et al^42^ and Yuan et al^43^ both examined microbiota of individuals with schizophrenia compared to healthy controls. Ma et al^42^ reported lower alpha diversity of chronically antipsychotic-treated schizophrenia (TSCZ) patients compared to first-episode drug-naïve schizophrenia (FSCZ) and healthy controls. In contrast, findings for beta diversity differed based on type of analysis. Unweighted UniFrac analysis demonstrated differences in beta diversity while weighted UniFrac showed no differences across groups. Differences in bacterial composition were also reported. Although there were no gut microbiota differences between FSCZ patients and healthy controls at the phyla level, the study reported eight differences at the family level, where the FSCZ patients had more Christensenellaceae, Enterobacteriaceae and less Victivallaceae, Pasteurellaceae, Turicibacteraceae, Peptostreptococcaceae, Veillonellaceae, Succinivibrionaceae than healthy controls. At the genus level, the study observed that the FSCZ patients had more *Escherichia* and less *Actinobacillus, Fusobacterium, Megasphaera*, and *SMB53* compared to healthy controls. Differences among the gut microbiota were seen among the TSCZ patients as well, where the relative abundances of the family Peptostreptococcaceae and Veillonellaceae and genus *Megasphaera, Fusobacterium*, and *SMB53* were increased compared to FSCZ patients. Lastly, TSCZ patients had higher levels of the family Enterococcaceae and Lactobacillaceae and genus *Escherichia, Enterococcus, Lactobacillus, Shigella, Streptococcus* and *Veillonella* compared to FSCZ patients, but these levels were similar among the FSCZ and healthy control groups. These findings suggest that antipsychotic treatment may affect some bacterial abundances but not others. These same bacteria were correlated with changes in the right middle frontal gyrus. Yuan et al^43^ study additionally had a 24-week risperidone intervention and concluded that the risperidone group had less *Bifidobacterium, Lactobacillus* and *Escherichia coli* and more *Clostriudium coccoides* at baseline compared to healthy controls. Following 24 weeks of treatment, *Bifidobacterium* and *E. coli* increased and *Lactobacillus* and *Clostridium coccoides* decreased. Metabolic parameters were also measured, and there were no correlations with changes in gut microbiota composition over the 24 week treatment period.

Lastly, Schwarz et al^44^ examined primary psychotic disorders and found that *Lactobacillus* was positively correlated with psychotic symptom severity. Schwarz et al^44^ examined the relationships between various bacterial groups with first episode psychosis (FEP). Their study found that the family Lactobacillaceae differed most among the FEP compared to healthy controls. *Lactobacillus* was positively correlated with the severity of psychotic symptoms. FEP patients with the strongest microbiota differences showed poorer response up to 12 months of antipsychotic treatment.

### ADHD

Only one study examined the gut microbiome of young adults with ADHD. While this study included many participants over 25 years of age in the healthy control group, the average age of the ADHD group was 19. Since this study by Aarts et al^45^ included 16s rRNA sequencing that included subsequent functional analysis, they were able to predict that the genus *Bifidobacterium* was responsible for the increase of cyclohexadienyl dehydratase (CDT) of ADHD participants. CDT is involved with the synthesis of phenylalanine, a dopamine precursor. The abundance of CDT in both ADHD and healthy groups was negatively correlated with bilateral ventral striatal differences for reward anticipation, as measured by fMRI.

## Discussion

Recent interest in the gut microbiome, the gut-brain-axis, and technologies to sequence and analyze data has paved the way for an increase in research to understand the links between the microbiome and mental health prevention and treatment. The scope of research in this field is broad, ranging from observational studies to clinical trials, child behavior to adult serious mental illness, and probiotic treatment to fecal transplantation. Since incidence and prevalence during earlier ages are linked to overall poorer health during adulthood, this review aimed to assess the breadth of microbiome research focused on child, adolescent, and young adult behavioral and mental health and to identify existing research gaps. We found 18 studies that evaluated relationships between the gut microbiome and behavioral and mental health within the child, adolescent, and young adult populations. There were no studies on only the adolescent age group, although four studies on children included individuals up to 16 years old and one young adult study included adolescents as young as 13 years old. While the studies used a wide range of surveys and questionnaires to measure different behavioral and mental health symptomology, the majority of studies utilized 16s rRNA sequencing for microbiota characterization.

All studies reported significant associations between the gut microbiome and behavior or mental health status and problems. However, there was minimal consensus regarding how behavioral and mental health problems were related to microbial diversity or composition, possibly due to the different behavioral and mental health problems examined across the various studies. Varied population characteristics, including age and sex, may also contribute to these differences. Additionally, microbial composition varies considerably by geographical location, where even regional differences have been reported to be the strongest phenotypic determinant of microbiome variability.^46^ However, in the following discussion, we attempt to synthesize what has been reported regarding key relationships of mental health problems with microbial composition and diversity, noting any differences found based on age of the youth.

### Relationships of Mental Health to Microbial Composition

Gut microbiota are composed of different bacteria species. In the following sections, we discuss any key findings for specific bacteria as they relate to mental health problems.

### Results by Age Group

Among children/adolescents, a few studies examined microbial composition of ADHD participants and healthy controls. Consistent with previous studies exploring the gut microbiota of healthy children/adolescents, four phyla were reported dominant among healthy controls: *Firmicutes, Bacteroidetes, Proteobacteria*, and *Actinobacteria*.^37^ The same study reported that these phyla were dominant in medication naïve ADHD participants. Due to these similarities among healthy participants with medication naïve ADHD participants, gut microbiota composition differences may vary at a lower taxonomic level rather than at the phylum level. However, findings with the family level and alpha and beta diversity were inconsistent among reviewed studies.

While no major similarities among bacterial types were reported in this child/adolescent age group, four child/adolescent studies reported findings on the genera *Bifidobacteria*, a gram positive anaerobic bacteria that is one of the major genera that resides in our Gl tract. *Bifidobacteria* are believed to exert positive health benefits and among the first microbes to colonize the infant GI tract ^47^. However, three of the four child/adolescent ADHD studies reported lack of associations between ADHD and *Bifidobacterium* levels.^28,36,37^ The last of the ADHD studies reported a trend toward a higher abundance of *Bifidobacterium* being correlated with fewer ADHD symptoms at baseline in a sample of boys, but lower levels of *Bifidobacterium* were associated with fewer ADHD symptoms after treatment with micronutrients ^35^. On the other hand, the abundance of *Bifidobacterium* was decreased among children with symptoms of depression, compared to those who were not depressed, in a sample of children with orthostatic hypotension.^33^ In sum, the direction of relationships between levels of *Bifidobacterium* and children’s symptoms in ADHD versus depression were opposite, indicating that there may be diverse ways in which bacteria are associated with different mental health problems. Although the general consensus has been that *Bifidobacteria* may be beneficial in the mental health or physical health of infants and adults ^48,49^, it may have different effects on children/adolescents and with different mental health conditions, as evident in the inconsistent findings from our literature review.

Similar to our findings with children/adolescents, there were few similarities among the young adult studies on the gut microbiome and mental health. Three studies reported findings on *Bifidobacterium* and *Lactobacillus*, with one reporting on both genera and the other two reporting on either one or the other. *Bifidobacterium* and *Lactobacillus* were lower in drug-naïve first-episode schizophrenia patients.^39^ *Bifidobacterium* increased but *Lactobacillus* decreased after risperidone treatment. In contrast, more *Lactobacillus* was correlated with psychotic symptom severity among a group of patients with primary psychotic disorders.^43^ Consistent with literature of adults, *Bifidobacterium* in these two studies was lower in participants with mental health conditions, supporting the view that more *Bifidobacterium* may be beneficial to the mental health of young adults. Results for *Lactobacillus* are less clear. *Lactobacillus* is another bacterium identified as beneficial in the literature^50^ and has been found in many functional foods and probiotics. Discrepant findings may be due to psychiatric diagnosis, differential receipt of treatment, body weight and body mass index, diets, and geographic regions as these factors have significant influences on the human microbiome.

### Internalizing symptoms

Nine studies examined the associations between composition of the gut microbiota and internalizing symptoms, six of which were on children/adolescents and three on young adults. Among the 9 studies examining the associations between gut microbiota and internalizing symptoms, the relationship of the abundance of *Bifidobacteria* with depressive symptoms was reported in Ishii et al’s^33^ study on children with orthostatic intolerance and Nishida et al’s^39^ study on young adults. Results from both studies reported similar findings that *Bifidobacteria* may play a beneficial role, reflected in its association with fewer internalizing symptoms. As discussed in the previous section about age, *Bifidobacteria* was decreased among depressed children, compared to those who were not depressed. The fact that both studies found the same results for *Bifidobacteria* is promising since there are 300-500 bacterial species harbored in our GI tract. Nonetheless, further studies are necessary to confirm these findings.

### Relationships of Mental Health to Microbial Diversity

Microbial diversity is a measure of the total number of bacterial species (richness) and the abundance of each type of organism. There is general consensus that more diverse microbiota may have a better capacity to withstand external threats and be associated with more optimal health.^51,52^ In this section, we highlight findings specific to diversity and mental health.

### Results by Age Group

Among the child/adolescent studies, several did not find any significant associations between alpha diversity or beta diversity with behavioral and mental health outcomes.^27,29,30,33,34,36^ ADHD was the primary mental health problem for which significant findings on diversity were reported. Consistent with previous research examining many aspects of health and disease, alpha diversity was decreased among young, male ADHD patients compared to healthy controls in Prehn-Kristensen’s et al’s^28^ study. In Wang et al’s^37^ research, there were mixed findings. Using the Simpson Index of Diversity, children with ADHD had lower diversity than healthy controls, a finding congruent with the previous study. However, Wang’s group found that ADHD patients had higher diversity than controls when using the Shannon and Chao Indices to measure diversity. One other study of Belgian primary school children found significant results.^32^ They reported that high stress was associated with lower alpha diversity, but specifically among preadolescents, not adolescents. Although there is no complete consistency, overall, study findings indicate that children with mental health challenges (ADHD or high stress) have less microbial diversity than children without them.

Results for young adults were inconsistent across studies in the relationship between mental health and microbial diversity. Two studies did not examine diversity,^43,44^ and findings in the other studies were for a two different mental health conditions (psychosis and ADHD). Higher alpha diversity was associated with major depression in Jiang et al’s^40^ study but not significantly associated with ADHD in Aarts et al’s^45^ study. In Kato-Kataoka et al’s^38^ study on risk for schizophrenia. In sum, in the younger age group, less alpha diversity was associated with worse mental health; whereas, greater alpha diversity may be associated with mental health problems in young adults.

### Internalizing Symptoms

Only three of the nine studies focused on internalizing symptoms reported significant findings for diversity. As compared to studies of middle and older adult age groups with depressive disorders, Jiang et al’s^40^ research on young adults reported an increase in alpha diversity among participants with A-MDD compared to healthy controls. On the other hand, high stress was associated with lower alpha diversity with differences among beta diversity between high and low stress groups.^32^ Similarly, alpha diversity was increased in medical students exposed to stressful situations.^38^ These disparate findings may be due to the different mental health conditions examined as well as age groups under study. Jiang et al’s^40^ study focused on two distinct groups, those diagnosed with a depressive disorder and healthy participants, whereas the two other studies measured symptoms of stress among healthy participants. Microbial diversity may be substantially different for individuals who have a diagnosis of a mental disorder and those who are generally healthy but experience psychological symptoms related to stressors in their life. Major differential factors for these groups are likely the chronicity and severity of their mental health challenges which may affect diversity over time.

### Additional Findings

All studies used the gut-brain-axis as a framework to guide their research aims. Pre-clinical studies have consistently shown bidirectional interactions within the gut-brain axis.^53^ While these reviewed studies have acknowledged the different pathways linking the gut, brain, and mental health, only four studies gathered biomarkers to measure the links of the gut microbiome with the brain or behavioral and mental health. Four studies reported cortisol levels, a biomarker often used to measure the hypothalamic-pituitary-adrenal (HPA) axis (endocrine pathway). Both studies by Michels et al^27,32^ on children reported no relationships between hair cortisol on SCFAs, fecal calprotectin, and gut microbiota composition. Nishida et al’s^39^ study reported null associations between probiotic administration and salivary cortisol levels while Kato-Kataoka et al’s^38^ study reported higher salivary cortisol levels in the placebo group only. Variations in methods of measurement (hair versus saliva), timing of collection (exposed to stressor or not), and small sample sizes may contribute to these differences in findings. Lastly, probiotics are hypothesized to alter the gut microbiota, but rather than probiotics having a direct effect on the HPA axis through cortisol measurement, they may have serotonergic effects via the neural pathway or immunomodulatory effects on the brain and behavior and mental health. **Strengths and limitations**

This is the first literature review of any kind examining the gut microbiota across age groups ranging from 2 to 25 years of age. However, the studies reviewed present several limitations. Half of these studies were cross-sectional, precluding our ability to identify the direction of associations between the gut microbiome and behavior/mental health. Some studies included self-reported or parent-reported measures rather than clinician-administered measures of behavior and mental health. Additionally, generalizability to clinical populations may be limited since some studies involved healthy participants, a specific subgroup of healthy participants (medical students), or medical patients in hospital settings. Nonetheless, measurement of severity of symptoms (a continuous variable) rather than diagnosis increases power to detect effects. On the other hand, studies that differentiate participants with severe illness via diagnosis from healthy controls can provide more insight to actual differences seen in the microbiome for various mental health groups. Confounds such as age, sex, medications, lifestyle factors, and diet were not always included in the statistical models and may therefore affect the results.^54^ However, sample sizes in most of the studies were small, precluding their ability to include too many covariates.

While most studies performed stool sample analyses using 16s rRNA, their examination of various regions of RNA and using different methods (16s rRNA sequencing compared to shotgun metagenomic sequencing) could have influenced results. Methods to measure the gut microbiome also varied among all studies, but 16s rRNA sequencing was used in the majority of studies. The 16S rRNA gene includes nine hypervariable regions (V1-V9). The region(s) examined in the reviewed studies varied (V1-V4), and some studies did not indicate which regions were sequenced. Bukin et al^55^ reported that the V2 and V3 regions have higher resolutions for genera and species compared to the V3-V4 regions. When considering the methodology for future research studies to estimate microbial community diversity and bacterial composition, the V2 and V3 regions should be considered for precision and higher resolution. It has been suggested that standardized methods be used across studies, from sample collection and storage to DNA extraction, PCR or library construction, sequencing, and bioinformatic analysis, to enable accurate comparison and synthesis of findings.^54^ We support this recommendation. Additionally, the two studies by Nishida et al^39^ and Kato-Kataoka et al^38^ had some positive findings, but the probiotic treatments consisted of different strains, making comparison of findings difficult. Due to the limited research in the area of the gut microbiome and behavioral and mental health in these age groups, our search was broad to include symptoms and disorders as independent variables, dependent variables, as well as covariates. However, due to the broad scope of symptoms and disorders, it was difficult to make comparisons across studies.

## Conclusions

Although inconsistent findings were reported across a number of studies in this review, we identified a few results where potential agreement is emerging in research to date. Regarding composition of the microbiome, *Bifidobacteria* does appear to be associated with more optimal states of mental health for both children/adolescents and young adults. Especially for depression, research indicated that youth with depression had lower levels of *Bifidobacteria* than healthy controls or those with fewer depressive symptoms. For diversity of the microbiome, findings were very mixed. There was some level of agreement that lower alpha diversity was present among children and adolescents with ADHD and high stress. However, these findings were opposite those for young adults who had higher alpha diversity when diagnosed with both ADHD and Major Depressive Disorder.

Our synthesis of existing literature to date is based on a limited body of research on the associations between the gut microbiome and behavioral and mental health of youth. The fact that there was substantial inconsistency across studies is likely due to the differences in mental health problems being examined, the populations in which the problems were being assessed, and the research designs or measures used. These limitations should be addressed in future research, with investigators attempting to replicate studies to confirm findings that have been reported. In addition, there is a need to examine lower taxonomic levels than those examined by studies in this review to determine if differences are apparent with more in depth assessment of the intestinal ecology. Given that many of the studies are cross-sectional, more longitudinal research would be helpful to assess for directionality. Clinical trials to include the effects of probiotics with the same strains and measurement of the gut microbiome are necessary for future recommendations of prevention, treatment, and management. Further in-depth sequencing and analyses such as shotgun metagenomic sequencing and fecal metabolomic analysis, as well as the inclusion of endocrine and inflammatory biomarkers, would be useful in clarifying the role of the gut microbiota in the gut-brain axis. Finally, utilizing the same metrics to measure and describe diversity of gut microbiota composition will allow for comparisons of mental health conditions and study populations.

## Data Availability

N/A. Data is already public.

---

**Figure 1.**
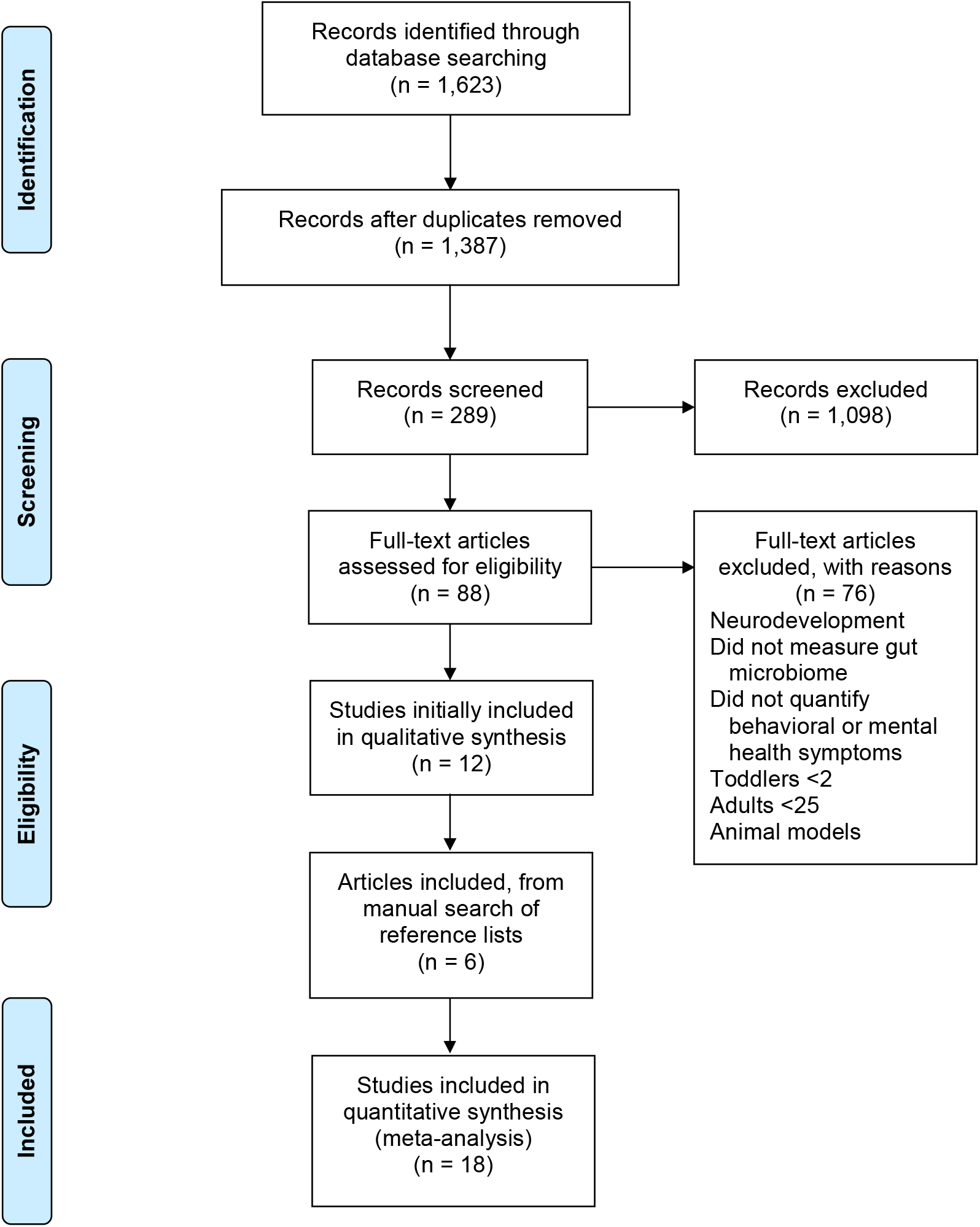
PRISMA flowchart showing the studies identified during the literature search and abstraction process

